# Calcium Channel Blockers: clinical outcome associations with reported pharmacogenetics variants in 32,000 patients

**DOI:** 10.1101/2022.03.23.22272805

**Authors:** Deniz Türkmen, Jane A.H. Masoli, João Delgado, Chia-Ling Kuo, Jack Bowden, David Melzer, Luke C. Pilling

**Author notes:** **Corresponding author** Dr Luke Pilling, College House (room 1.09), Epidemiology and Public Health Group, University of Exeter, Magdalen road, St. Luke’s Campus, Exeter, EX1 2LU, UK.

## Abstract

**Background:** Dihydropiridine calcium channel blockers (dCCB) (e.g. amlodipine) are widely used for treating hypertension. Pharmacogenetic variants impact treatment efficacy, yet evidence on clinical outcomes in routine primary care is limited. We aimed to estimate associations between reported pharmacogenetic variants and incident adverse events in a community-based cohort prescribed dCCB, including in high-risk subgroups.

**Methods:** We analysed up to 32,360 UK Biobank European-ancestry participants prescribed dCCB in primary care electronic health records (from UK General Practices, 1990 to 2017). We investigated 23 genetic variants in 16 genes reported in PharmGKB, including *CYP3A5* and *RYR3*. Outcomes were incident diagnosis of coronary heart disease (CHD), heart failure (HF), chronic kidney disease (CKD), edema, and switching antihypertensive medication. Secondary analysis in patients with history of heart disease was also performed.

**Results:** Participants were aged 40 to 79 years at first dihydropyridine prescription (treatment duration 1 month to 40 years, mean 5.9 years). Carriers of rs877087 T allele in the ryanodine receptor 3 (*RYR3*) had increased risk of HF (Hazard Ratio 1.13: 95% Confidence Intervals 1.02 to 1.25, p=0.02). We estimated that if rs877087 T allele carriers were prescribed an alternative treatment the incidence of HF in patients prescribed dCCB would reduce by 9.2% (95%CI 3.1 to 15.4). In patients with a history of heart disease when first prescribed dCCB (N=2,296), *RYR3* rs877087 homozygotes had increased risk of new CHD or HF compared to CC variant (HR 1.25, 95%CI 1.09 to 1.44, *p*=0.002). Two variants increased likelihood of switching to an alternate antihypertensive medication (rs10898815 in gene *NUMA1* HR 1.16: 95%CI 1.07 to 1,27, p=0.0009; rs776746 in *CYP3A5* HR 1.59: 95%CI 1.09 to 2.32, p=0.02). rs776746 in *CYP3A5* also increased CKD risk (HR 2.12, p=0.002). The remaining previously reported variants were not strongly or consistently associated with the studied clinical outcomes.

**Conclusions:** In this large primary care cohort, patients with common genetic variants in *NUMA1, CYP3A5* and *RYR3* had increased adverse clinical outcomes. Work is needed to establish whether outcomes of dCCB prescribing could be improved by prior knowledge of such pharmacogenetics variants supported by clinical evidence of association with adverse events.

## Introduction

High blood pressure - hypertension - is a key modifiable risk factor for cardiovascular morbidity and mortality. While reducing raised blood pressures is the goal, only one-third of hypertensive patients treated with antihypertensive medications are estimated to reach target blood pressures (1). The reasons for failure to control raised blood pressure are complex, but genetic factors are proposed to play a role, either directly on blood pressure or indirectly by influencing antihypertensive medication response, adverse events or medication adherence(2).

Calcium channel blockers (CCB) are the first line recommended antihypertensive for most adults with hypertension, and their use is widespread across the world (3–5). There are two subgroups of CCBs, the most common being Dihydropyridines (dCCB), which are regarded as relatively safe and cost-effective (6). Edema is a common dCCB adverse effect, with incidence rates of 22% (7–9), that affects the quality of life of patients and can lead to discontinuation of treatment (10,11). The presence of edema can result in additional prescribing, which in turn can cause additional adverse outcomes including falls, overdiuresis, acute kidney injury and polypharmacy (11,12). Genetic factors can predispose side effects, as well as further complications.

The pharmacogenomics knowledge base (PharmGKB) documents genetic variants reported to influence dCCB effectiveness or adverse events (2,13–15). The levels of supporting evidence for each variant is variable, with many only having limited clinical evidence. Such evidence includes: Reported genes containing single-nucleotide polymorphisms (SNPs) include those encoding calcium channel subunits themselves, such as the voltage-gated calcium channels α1C (*CACNA1C*). SNPs in other ion-channels which are reported to alter dCCB responses (including *PICALM, TANC2, NUMA1, APCDD1, GNB3, SLC14A2, ADRA1A, ADRB2*, and *CYP3A4*). SNPs in ATP-binding cassette subfamily B member 1 (*ABCB1*) and in cytochrome p450 3A5 (*CYP3A5*) are reported to affect the clearance of dCCBs, and SNPs in cytochrome p450 oxireductase *(POR*) reportedly influence the plasma concentration of medicines. SNPs in nitric oxide synthase 1 adaptor protein (*NOS1AP*) increase risk of cardiovascular death, And SNPs in ryanodine receptor 3 *(RYR3)* and in atrial natriuretic precursor A (*NPPA*), are reported to increase the risk of cardiovascular disease. Particularly, *RYR3* (in intracellular calcium channels) was found to be associated with heart failure (HF) and it is a need to examine its effect on stroke, and effects on heart disease in risky groups as it is unknown (16).

Evidence of impact on clinical outcomes, especially in routine primary care (rather than in acute hospital care settings) is currently limited for most pharmacogenetics variants reported to affect dCCBs. Here we analyse the UK Biobank (UKB) community volunteer cohort with linked genetic and medical records. We aimed to determine the extent to which 23 commonly occurring (minor allele frequency >3%) pharmacogenetic variants in 16 genes reported to affect dCCB effectiveness or rates of adverse events - are associated with clinical outcomes.

## Methods

### UK Biobank cohort

The UK Biobank enrolled 503,325 community-based volunteers aged 40-70 years who visited one of 22 assessment centres in Wales, Scotland, or England in 2006-2010 (17). Extensive questionnaires on demographic, lifestyle and health information data were collected at the baseline assessment. Blood samples for genetic and biochemical analyses, and anthropometric measures were gathered. This study of dihydropyridines was conducted using the linked GP (primary care) data available in 230,096 participants. Data were available between Jan 1990 and August 2017 (see below for details).

### Ethics

Participants gave consent to receive relevant information about clinical findings at baseline only: therefore, UKB data on individual genetic status were not reported to participants or their clinicians and could not therefore have influenced prescribing. The Northwest Multi-Centre Research Ethics Committee approved the collection and use of UK Biobank data (Research Ethics Committee reference 11/NW/0382).

### General Practice (GP) data

More than 57 million prescriptions for 230,096 (45.7%) participants in the primary care data were recorded. The GP data was available up to 31 May 2016 (England TPP system supplier) and 31 August 2017 (Wales EMIS/Vision system). Drug name, quantity, date of prescription, drug code (in clinical Read v2, British National Formulary (BNF), or dm+d (Dictionary of Medicines and Devices) format, depending on suppler) are available. We identified prescribing records for all antihypertensive medications. dCCB CCBs (amlodipine: Norvasc®, Katerzia®, Istin®, Amlodipine®; felodipine: Plendil®, Cardioplen®, Folpik®, Felotens®, Vascalpha®, Neofel®, Parmid®; lacidipine: Lacipil®, Motens®; Molap®, Lacidipine®; nisoldipine: Sular®, lercanidipine: Zanidip®, Lercanidipine®, nimodipine: Nimotop®, Nymalize®, nifedipine: Adalat®, Afeditab®, Procardia®, Adipine®, Nifedipress®, Tensipine®, Dexipress®, Valni®, Adanif®, Neozipine®, Nidef®, Nifedipine®, Fortipine®, Coracten® ; nitrendipine: Baypress®, nicardipine: Cardene®) examined in more detail with the date first prescribed, date last prescribed, number of total prescriptions, and average number of prescriptions over the treatment span.

Patients with a date of last prescription at least 3 months prior to the end of available GP data were defined as having discontinued treatment (we chose 3 months because it is rare for patients to receive more than 3 months of medications in a single prescription). The end date was either the date of deduction (removal from GP list, where available) or 31 May 2016 where no deduction date was present (i.e., still registered at an available practice). Data after 31 May 2016 is incomplete, depending on GP provider (see UK Biobank documentation (18)).

We also identified antihypertensive prescriptions apart from dCCB; diuretics, beta blockers, alpha blockers, angiotensin converting enzyme inhibitors (ACEIs) and other antihypertensives using the Read 2 codes and BNF codes.

### Disease ascertainment

Primary and secondary care health records were used to examine the dCCB related adverse events. Edema diagnosis was ascertained from ICD-10 and ICD-9 codes(19) :we included ICD-10 codes (R60.0 Localized oedema, R60.1 Generalized oedema, R60.9 Oedema, unspecified) and ICD-9 codes (7570, 7823, 2766) for edema. We converted these to Read codes (C3661, C3662, M2A.., R023., PH0.., PH00., PH01., PH02., PH0z., R023., C366.) used in UK primary care records using UK Biobank-provided diagnostic code maps.

Cardiovascular events from hospital admissions records were available up to 14 years follow-up after baseline assessment (HES in England up to 30 September 2020: data from Scotland and Wales censored to 31 August 2020 and 28 Feb 2018, respectively), covering the entire period up to the date of censoring of primary care prescribing data. Diagnosis of MI/angina was ascertained using ICD-10 codes I20*, I21*, I22*, I23*, I24* and I25*; ischemic stroke was using ICD-10 codes I63*; chronic kidney disease (CKD) using ICD10 codes N18* and Y84.1; and heart failure using ICD10 codes I50* and J81*.

### Genetic Variants

Directly genotyped genetic variants (n=805,426) were ascertained in 488,377 UK Biobank participants, which used two almost identical platforms sharing >95% of variants: the Affymetrix Axiom UKB array (in 438,427 participants) and the Affymetrix UKBiLEVE array (in 49,950 participants). Extensive quality control was applied by the central UK Biobank team (21). Genotype imputation increased the number of genetic variants to ∼96 million in 487,442 participants (21). Our analysis included 451,367 participants (93%) identified as genetically European (identified by genetic clustering, as described previously (22)): unfortunately, sample sizes from other ancestry groups were too small to analyse separately.

We analysed the genetic variants with documented effects on dCCBs effectiveness in the literature (23) and in the PharmGKB database. This included 29 SNPs in the following genes: *NPPA, NOS1AP, CYP3A4, GNB3, RYR3, CACNA1C, ABCB1, ADRA1A, SLC14A2, ADRB2, POR, PICALM, TANC2, NUMA1, APCDD1* (see Supplementary Table 1 for details). Genotype status for 23 variants could be ascertained from the available UK Biobank imputed data (release version 3) and minor allele frequencies were common enough to study (frequency varying from 3% to 46% - see Supplementary Table 1 for details) in the UKB cohort: results for all 23 studied SNPs are reported.

### Primary Analysis

Associations between genotypes and outcomes (GP-diagnosed edema, and hospital-diagnosed CHD (MI/angina), HF, and CKD) were estimated using Cox proportional hazards regression models. They were adjusted for age at first prescription, sex, and genetic principal components 1 to 10 (to adjust for population genetic ancestry) in patients who were prescribed dCCBs by their GPs (General Practice). We included patients if they have had at least two prescriptions and more than 2 months of dCCBs treatment. Patients exited the model on the hospital inpatient record censoring dates i.e., for cardiovascular and renal outcomes we therefore performed an ‘intention to treat’ analysis, reducing any bias due to discontinuation before diagnosis of adverse outcomes (e.g., discontinuing dCCB treatment and having heart diseases as a result). For the analysis of edema participants exited the model on the date of their last known prescription plus 2 months (the wash out period), as edema symptoms are alleviated after stopping medication. Unless otherwise stated all analyses were performed in STATA v16.

To estimate the *Genetically Moderated Treatment Effect* (GMTE) we used “TWIST” (Triangulation with a Study) (24), a novel pharmacogenetic causal inference approach. This enables estimation of the predicted outcome if all participants were reassigned the low-risk genotype, therefore providing an estimate of the genetic effect. In brief, the methods uses Aalen additive hazards regression models to test several assumptions common to pharmacogenetic analysis; primarily that the genetic variants do not predict whether an individual receives dCCB treatment; are not associated with any measured confounders predicting dCCB use or the studied outcome; and only affect the outcome through the interaction with dCCBs (see (24) for details). From this analysis the most efficient and robust estimate of the GMTE, is derived. Of note, the GMTE estimate may be the result of applying a single method, or instead be the combination of two or more estimates from different methods. We used R version 4.0.2 and R package ‘twistR‘ (https://github.com/lukepilling/twistR) v.0.1.3.

We also investigated the association between genotype and likelihood of switching dCCB for an alternative antihypertensive prescription. The association between genotypes and changing treatment was tested using Cox’s proportional hazards regression models, with adjustment for age at first prescription, sex, and genotyping principal components of ancestry 1-10 in patients. We considered the deduction date provided i.e., if patients were removed from a GP practice included in the UK Biobank data this became their date of censoring. We included patients if they had 3 months of GP data after their last known prescription (i.e., we can be more certain of a discontinuation of dCCB treatment rather than the last prescription being too close to the date for censoring).

To adjust for multiple statistical testing and control the false discovery rate we applied Benjamini-Hochberg correction to p-values for the associations between 23 SNPs and each outcome (using R function ‘p.adjust()‘). This approach is likely conservative, as it does not account for the prior evidence for the associations between these 23 SNPs and adverse events in patients prescribed dCCBs.

### Secondary Analysis in Patients with Heart Disease Diagnosis Prior to dCCB Treatment

We only included patients who had any heart diseases prior to the dCCB treatment for MI/angina/HF outcomes models as secondary analysis, as worsening angina and acute MI are reported as a caution for patients with coronary arteria disease by the Food and Drug Administration (FDA) in the prescribing information (9). We also tested associations for stroke and the *RYR3* calcium channel gene variant in patients on dCCBs, to examine the treatment effect, as stroke was associated with RYR3 in a GWAS (Genome Wide Association Study) regardless of use of dCCBs (25).

## Results

### Characteristics of the sample

There were 32,360 (45.6% female) patients who were prescribed dCCB CCB in primary care. The mean age was 61.3 (SD 7.7). The number of prescriptions in a year varied from 1 to 25, with a mean of 9.2 (SD 4.6). The mean prescription period was 5.9 (SD 5.2) years, the median was 4.4 (IQR 1.6 to 9.1). (See table 1 for details). The allele frequencies for the 22 studied genetic variants range from 3% to 50% (for details see Supplementary Table 2).

**Table 1.** Descriptive table of the UK Biobank participants included in the analysis.

| **GP prescribed** |  |
| --- | --- |
| Number of participants | 32,360 |
| Females, n (%) | 17,590(45,6) |
| Age at first prescription |  |
| Minimum : maximum | 40:79.3 |
| Mean (SD) | 61.3(7.7) |
| Number of prescriptions in a year |  |
| Minimum : maximum | 1:25 |
| Mean (SD) | 9.2(4.6) |
| Years between first and last prescription |  |
| Minimum : maximum | 0.08:39.9 |
| Mean (SD) | 5.9(5.2) |
| Edema^ pre prescriptions of dCCBs | 223(0.6) |
| MI/angina^^ pre prescription of dCCBs | 3,026(7.7) |
| CKD^^ pre prescriptions of dCCBs | 229(0.6) |
| Heart failure^^ pre prescriptions of dCCBs | 333(0.8) |
| Stroke^^ pre prescription of dCCBs | 333(0.8) |
| Prescribed other antihypertensive | 6,146 (15.6) |
| Changing treatment from dCCBs to others | 5,565 (14.2) |
| Edema^ post prescriptions of dCCBs | 400 (1.0) |
| MI/angina^^ post prescription of dCCBs | 7,430 (18.9) |
| CKD^^ post prescriptions of dCCBs | 2,940 (7.5) |
| Heart failure^^ post prescriptions of dCCBs | 2,292(5.8) |
| Stroke^^ post prescription of dCCBs | 1,001(2.6) |

*Analysis of European-ancestry participants with >1 dCCB prescription in the available GP prescribing data. Participants excluded if dCCB prescribing frequency was less than once every 2 months.*
*^Primary care recorded*
*^^Hospital diagnosed diseased*
*CKD: Chronic kidney disease*

#### RYR3

The ryanodine receptor 3 (*RYR3*) rs8777087 T allele prevalence in people on dCCB treatment in UK Biobank was 46%, and TT homozygotes was 21.3%.

2,292 of the 32,360 patients prescribed dCCBs developed heart failure during the follow up period. Diagnoses were more common in *RYR3* rs8777087 TT homozygotes (n=404, 6.1% of 6,607) and CT heterozygotes (n=943, 6.1% of 15,377) compared to common CC homozygotes (n=491, 5.4% of 9,090) (Figure 1; Table 2; see Supplementary table3 for details). The increased risk of hospital diagnosed heart failure was significant in Cox’s proportional hazards regression models adjusted for age, sex, and genetic ancestry (HR_TT vs CC_ 1.15, 95% CI 1.01 to 1.31, *p=*0.04 and HR_CT vs CC_ 1.12, 95% CI 1.01 to 1.25, *p*=0.04). Though results were not significant after Benjamini-Hochberg adjustment for multiple statistical testing (adjusted p>0.05) the burden of prior evidence increases plausibility of the associations. Heterozygotes (11.5%) were also more likely to have incident MI, angina, or HF compared to common CC homozygotes (10.3%) (HR 1.12, 95% CI 1.03 to 1.21, *p*=0.007) (Table 3). Heterozygotes were more likely to have incident stroke compared to CC homozygotes (HR 1.22, 95% CI 1.04 to 1.45, *p*=0.02) (Table 3).

**Figure 1.**
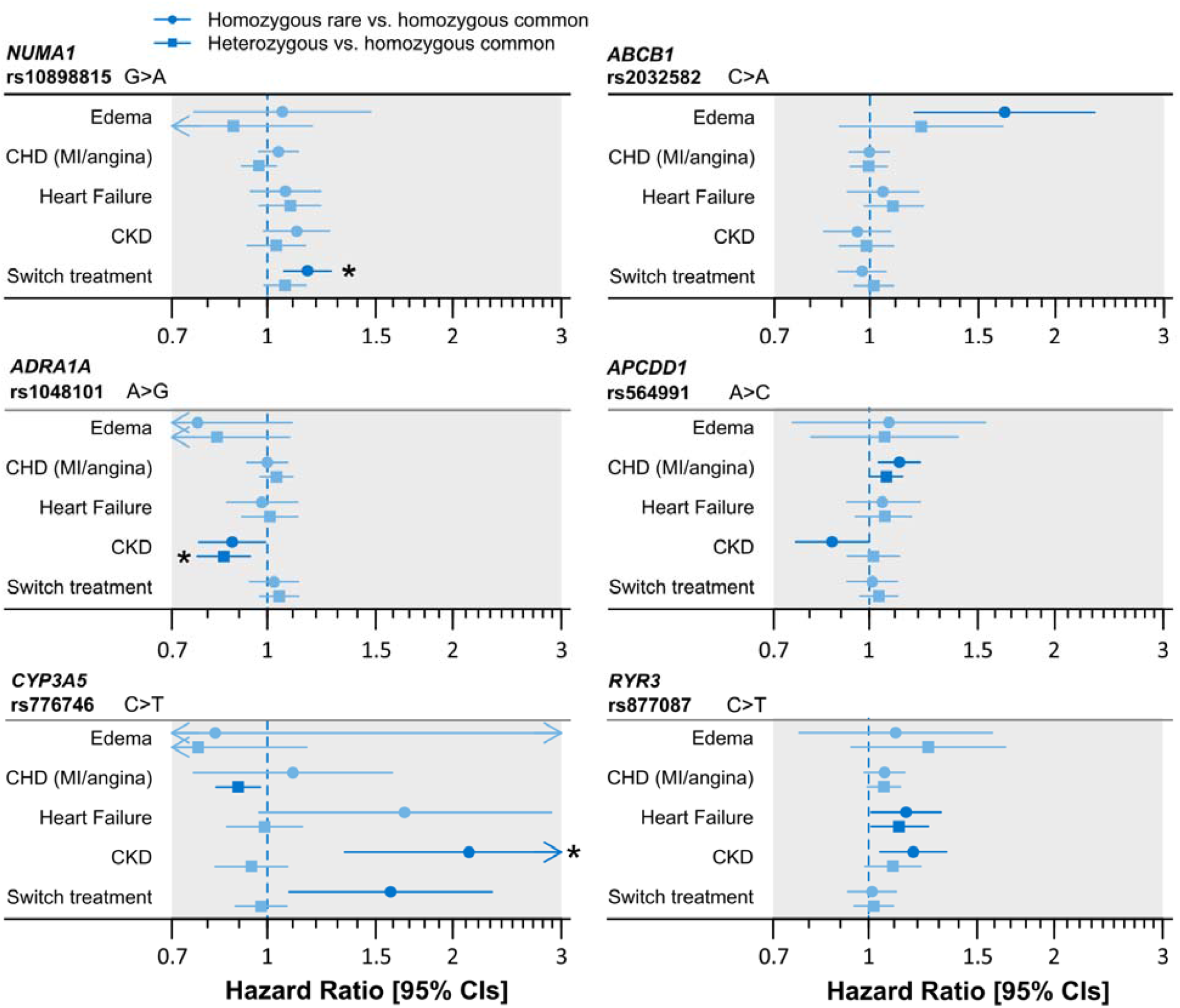
Associations between top six genotypes and dCCB-related adverse events. Six (of 23) variants with the most robust associations with clinical outcomes. For each genotype the association with 4 adverse event diagnoses (edema, CHD, heart failure, and CKD) and with likelihood of switching to an alternative antihypertensive treatment are shown for both heterozygotes and homozygotes, compared to the homozygous common genotype. Nominally significant associations (p<0.05) are indicated with darker colour, and associations significant after multiple testing correction (false discover rate, FDR) are indicated with stars. See Table 2 for details including numbers of heterozygotes/homozygotes and adverse events, and Supplementary Table 2 for results for all 23 genetic variants.

**Table 2.** Significant associations between genotypes and dCCB related adverse events.

| Outcome | Gene | Allele | N | n | % | HR | 95%CI |  | p |
| --- | --- | --- | --- | --- | --- | --- | --- | --- | --- |
| **Switching treatment** | *NUMA1*<br>rs10898815 | GG | 8,214 | 932 | 11.4 |  |  |  |  |
|  |  | GA | 16,104 | 1,940 | 12.1 | 1.07 | 0.99 | 1.16 | 0.10 |
|  |  | AA | 7,995 | 1,044 | 13.1 | 1.16 | 1.06 | 1.27 | 0.001 |
|  |  |  | 32,313 | 3,916 |  |  |  |  |  |
|  | *CYP3A5*<br>rs776746 | CC | 28,184 | 3,420 | 12.1 |  |  |  |  |
|  |  | CT | 4,024 | 476 | 11.8 | 0.98 | 0.89 | 1.08 | 0.64 |
|  |  | TT | 152 | 27 | 17.8 | 1.59 | 1.09 | 2.32 | 0.02 |
|  |  |  | 32,360 | 3,923 |  |  |  |  |  |
| **CKD** | *CYP3A5*<br>rs776746 | CC | 25,177 | 1,757 | 7.0 |  |  |  |  |
|  |  | CT | 3,545 | 239 | 6.7 | 0.94 | 0.82 | 1.08 | 0.39 |
|  |  | TT | 140 | 18 | 12.9 | 2.12 | 1.34 | 3.38 | 0.002 |
|  |  |  | 28,862 | 2,014 |  |  |  |  |  |
|  | *RYR3*<br>rs877087 | CC | 8,467 | 545 | 6.4 |  |  |  |  |
|  |  | TC | 14,231 | 1,008 | 7.1 | 1.10 | 0.99 | 1.22 | 0.09 |
|  |  | TT | 6,164 | 461 | 7.5 | 1.18 | 1.04 | 1.34 | 0.01 |
|  |  |  | 28,862 | 2,014 |  |  |  |  |  |
|  | *APCDD1*<br>rs564991 | AA | 9,709 | 675 | 7.0 |  |  |  |  |
|  |  | AC | 14,179 | 1,027 | 7.2 | 1.02 | 0.92 | 1.12 | 0.76 |
|  |  | CC | 4,910 | 306 | 6.2 | 0.87 | 0.76 | 1.00 | 0.04 |
|  |  |  | 28,798 | 2,008 |  |  |  |  |  |
|  | *ADRA1A*<br>rs1048101 | AA | 8,751 | 672 | 7.7 |  |  |  |  |
|  |  | AG | 14,277 | 947 | 6.6 | 0.85 | 0.77 | 0.94 | 0.001 |
|  |  | GG | 5,834 | 395 | 6.8 | 0.88 | 0.77 | 0.99 | 0.04 |
|  |  |  | 28,862 | 2,014 |  |  |  |  |  |
| **HF** | *RYR3*<br>rs877087 | CC | 9,090 | 491 | 5.4 |  |  |  |  |
|  |  | CT | 15,377 | 943 | 6.1 | 1.12 | 1.01 | 1.25 | 0.040 |
|  |  | TT | 6,607 | 404 | 6.1 | 1.15 | 1.01 | 1.31 | 0.04 |
|  |  |  | 31,074 | 1,838 |  |  |  |  |  |
| **CHD** | *APCDD1*<br>rs564991 | AA | 10,299 | 1770 | 17.2 |  |  |  |  |
|  |  | AC | 15,111 | 2771 | 18.3 | 1.07 | 1.00 | 1.13 | 0.04 |
|  |  | CC | 5,262 | 1004 | 19.1 | 1.12 | 1.04 | 1.21 | 0.004 |
|  |  |  | 30,672 | 5,545 |  |  |  |  |  |
| **Edema** | *ABCB1*<br>rs2032582 | CC | 8,956 | 62 | 0.7 |  |  |  |  |
|  |  | AC | 14,866 | 125 | 0.8 | 1.21 | 0.89 | 1.64 | 0.22 |
|  |  | AA | 6,354 | 75 | 1.2 | 1.65 | 1.18 | 2.32 | 0.003 |
|  |  |  | 30,176 | 262 |  |  |  |  |  |

N = total number of genotype group in analysis. n = number of diagnoses. %=percent of genotype group with a diagnosis.

**Table 3.**
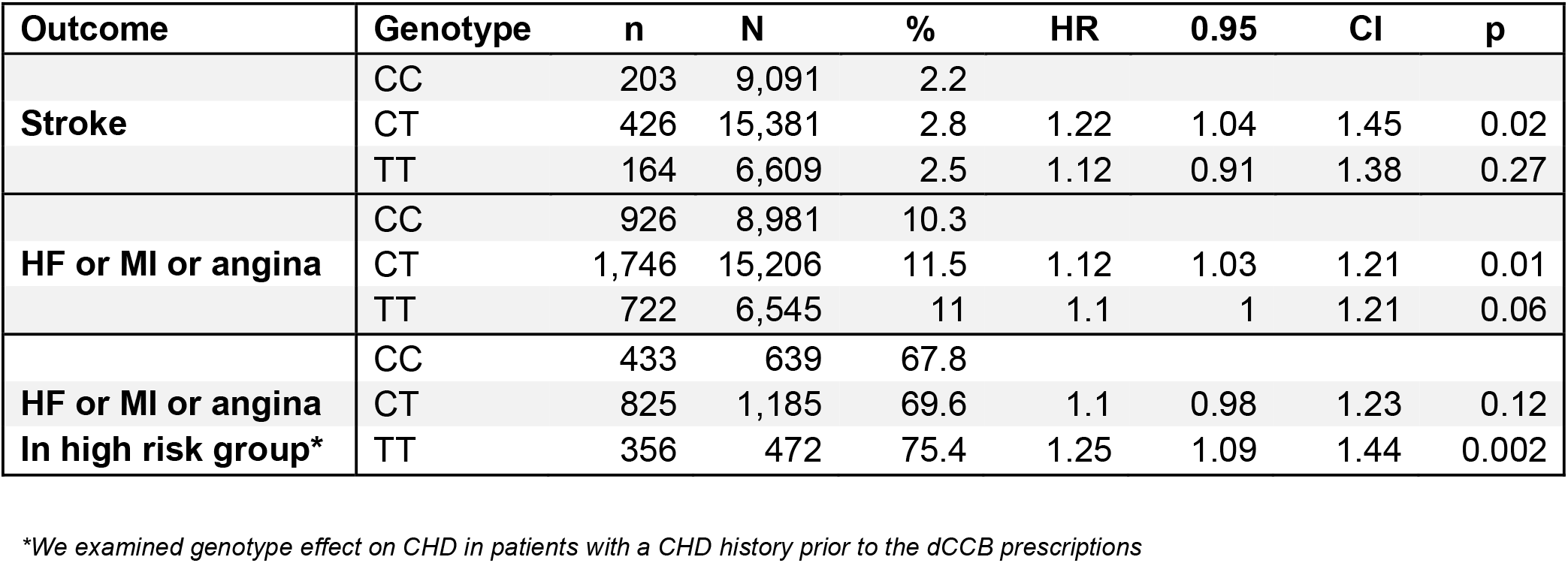
Associations between hospital-diagnosed CHD and stroke and variants of *RYR3* in patients with CHD diagnosed prior to dCCB prescription.

We used the TWIST (24) framework to estimate that the overall incidence of heart failure in patients prescribed dCCBs could be reduced by 9.2% (95%CI 3.1 to 15.4) if rs877087 T allele carriers received the same treatment benefit as non-carriers i.e. were switched to an alternative antihypertensive medication unaffected by rs877087 genotype.

To give further details on the TWIST results: because the association with HF was similar between heterozygotes and minor allele homozygotes we estimated the GMTE in carriers (any rs877087 T allele) compared to CC homozygotes. rs8777087 was not associated with HF in individuals never prescribed dCCBs (GMTE0 estimate p>0.05, Supplementary Table 5). From TWIST we found the ‘robust’ GMTE (RGMTE) and the ‘Mendelian randomization’ estimates could be combined to give a more efficient and precise estimate. The risk of HF was 0.069% greater per year after treatment initiation in carriers compared to non-carriers (p=0.003; Supplementary Table 4). When multiplied by the number of genotype-carrier patient-years in the model (244,818) and divided by the total number of diagnoses in the treated individual (1,838) we estimate that if carriers of the rs877087 T allele could experience the same treatment effect as non-carriers, 170 HF diagnoses could have been avoided (95%CI 58 to 282), hence the 9.2% quoted earlier.

In the subgroup of patients with a pre-existing heart disease (MI, angina, or HF) at the start of dCCB prescribing, *RYR3* TT homozygotes had an increased risk of developing incident heart diseases compared to common CC homozygotes (75.4% vs 67.8) with a HR 1.25 (95% CI 1.09 to 1.44, *p*=0.002) (Table 3; See Supplementary table 5).

Overall, 2,940 (7.5%) patients on dCCBs had incident chronic kidney disease (CKD). *RYR3* rs877087 TT homozygotes (461 CKD cases in 6,164 TT homozygotes) were more likely to have hospital diagnosed CKD compared to the common homozygotes groups (HR 1.18, 95% CI 1.04 to 1.34, *p*=0.01) (See supplementary table 6 for details). TWIST results showed that rs877087 was not associated with CKD in individuals never prescribed dCCBs (GMTE0 estimate p>0.05, Supplementary Table 4). The risk of CKD was 0.082% (p=0.002) greater per year after treatment in carriers compared to non-carriers (Supplementary Table 4). We estimate that if carriers of the rs877087 T allele could experience the same treatment effect as non-carriers 199 CKD diagnoses could have been avoided (95%CI 75 to 324). Therefore, the overall incidence of CKD in patients prescribed dCCBs could be reduced by 8.6% if rs877087 T allele carriers received the same treatment benefit as non-carriers (95%CI 3.2 to 14.0).

#### CYP3A5

Patients with *CYP3A5* rs776746 TT (CYP3A5*3) genotype (0.47% of patients), a variant previously linked to kidney related outcomes (26) had increased risk of CKD (HR 2.12: 95%CI 1.34 to 3.38, *p*=0.002) compared to GG homozygotes (Figure 1; Table 2). The association was still significant after Benjamini-Hochberg adjustment for multiple statistical testing (adjusted p<0.05). When we repeated the analysis for patients who were on dCCB but had no CKD history, 12.3% of *CYP3A5* rs776746 TT homozygotes without prevalent CKD were diagnosed with incident CKD compared to 6.6% of heterozygotes and 6.8% homozygotes for CC, (HR 2.09, 95% CI 1.29 to 3.37, *p*=0.003) (See supplementary table 7 for details). TWIST results showed that rs776746 was not associated with CKD in individuals never prescribed dCCBs (GMTE0 estimate p>0.05, Supplementary Table 4). The risk of CKD was 0.69% (p=0.003) greater per year after treatment in rs776746 TT homozygotes compared to CC homozygotes (Supplementary Table 4). We estimated that if rs776746 TT homozygotes could experience the same treatment effect as CC homozygotes 11 CKD diagnoses could have been avoided (95%CI 4 to 18). Therefore, the overall incidence of CKD in patients prescribed dCCBs could be reduced by 0.5% (95%CI 0.2 to 0.9) if rs776746 TT homozygotes received the same treatment benefit as CC homozygotes.

Of the patients on dCCB prescription, 5,565 (14.2%) changed treatment from dCCB CCBs to other antihypertensives. *CYP3A5* rs776746 TT homozygotes (n=27/152) were also more likely to change treatments compared to common homozygotes; HR 1.59, 95% CI 1.09 to 2.32, *p*=0.02, respectively (See Figure 1; Table 2; See supplementary table 8 for details).

#### NUMA1

Of those 5,565 patients switched treatment, 1,044 were *NUMA1* rs10898815 AA homozygotes (n=7,995). AA homozygotes were more likely to switch treatments compare to their common homozygotes (HR 1.16, 95% CI 1.06 to 1.27, *p=*0.001) (Figure 1; Table 2; See Supplementary table 8 for details). The association was still significant after Benjamini-Hochberg adjustment for multiple statistical testing (adjusted p<0.05).

#### ADRA1A

Adrenoceptor Alpha 1A *(ADRA1A)* rs1048101 AA homozygotes had an increased risk for CKD (HR 1.18, 95% CI 1.04 to 1.34, *p*=0.01) compared to GG homozygotes (Figure 1; Table 2). GG homozygotes (395 cases of 5,834 patients) and AG heterozygotes (947 cases of 14,277 patients) were associated with decreased risk of CKD compared to AA homozygotes (HR 0.88, 95% CI 0.77 to 0.99, p=0.04 and HR 0.85, 95%CI 0.77 to 0.94, p=0.001, respectively). The association was still significant after Benjamini-Hochberg adjustment for multiple statistical testing (adjusted p<0.05). TWIST analysis showed that rs1048101 was not associated with CKD in individuals never prescribed dCCBs (GMTE0 estimate p>0.05, Supplementary Table 4). The risk of CKD was 0.08% (p=0.02) greater per year after treatment in AA homozygotes compared to GG homozygotes (Supplementary Table 4). We estimated that if rs1048101 AA homozygotes could experience the same treatment effect as non-carriers (i.e. were prescribed an alternative antihypertensive medication unaffected by this genotype) 86 CKD diagnoses could have been avoided (95%CI 13 to 138). Therefore, the overall incidence of CKD in patients prescribed dCCBs could be reduced by 7% (95%CI 1.1 to 12.9) if rs1048101 AA homozygotes received the same treatment benefit as GG homozygotes.

#### APCDD1

Of those patients on dCCB prescriptions, 7,430 (18.9%) patients had incident MI/angina post dCCB treatment. Of those 7,430 patients, 1,004 was homozygotes for *APCDD1* rs564991 CC. 19.1% homozygotes for *APCDD1* rs564991 CC had the increased risk for MI/angina compared to 18.3% heterozygotes and 17.2% homozygotes for AA, (HR 1.12, 95% CI 1.04 to 1.21, *p=*0.004 for CC and HR 1.07, 95% CI 1 to 1.13, *p*=0.04 for AC) (Figure 1; Table 2; See Supplementary table 9 for details). TWIST analysis showed that rs564991 was not associated with CHD in individuals never prescribed dCCBs (GMTE0 estimate p>0.05, Supplementary Table 4). The risk of CHD was 0.19% (p=0.002) greater per year after treatment in CC homozygotes compared to AA homozygotes (Supplementary Table 4). We estimated that if rs564991 CC homozygotes could experience the same treatment effect as AA homozygotes 98 CHD diagnoses could have been avoided (95%CI 35 to 162). Therefore, the overall incidence of CHD in patients prescribed dCCBs could be reduced by 3.5% (1.3 to 5.8) if rs564991 CC homozygotes received the same treatment benefit as AA homozygotes.

306 patients of 4,910 *APCDD1* rs564991 CC homozygotes had CKD after dCCB treatment. They were less likely to have CKD compared to their common homozygotes (6.2% vs 7%) (HR 0.87, 95% CI 0.76 to 1, *p*=0.04). However, these were not significant after adjusting for multiple statistical testing. TWIST analysis estimated that is rs564991 CC homozygotes experienced the same treatment effect as AA homozygotes (i.e. were switched to an alternative antihypertensive) this may increase the overall CKD incidence in patients prescribed dCCBs by 63 diagnoses (95%CI 23 to 104, p=0.002).

#### ABCB1

Only 400 (1.02%) patients were recorded as having edema in their GP records, after dCCB treatment. ATP-binding cassette subfamily B member 1 (*ABCB1*) rs2032582 AA homozygotes were one of the two variants associated with edema (Supplementary table 10 for details). AA homozygotes were more likely to have edema with a rate of 1.18% (n=75/6,354, HR 1.65, 95% CI 1.18 to 2.32, *p*=0.03) compared to common CC homozygotes with a rate of 0.69% (n=62/8,956) (Figure 1; Table 2).

### Other variants in genes

There were additional associations with specific outcomes for several other variants, although as these were based on very small numbers or were combined with contrary results between heterozygotes and homozygotes, these results should be treated with caution.

Patients heterozygotes for G protein subunit beta 3 *(GNB3)* rs5443 (n=2,434/13,017) had an increased risk for MI/angina compared to homozygotes for CC (HR 1.07: 95%CI 1.01 to 1.13, *p*=0.02). Incident MI/angina was less likely to occur in patients heterozygous for *CYP3A4* rs2740574, *CYP3A5* rs776746 and Nitric oxide synthase 1 adaptor protein (*NOS1AP)* rs12143842 (*p*=0.04, *p*=0.01 and *p*=0.03, respectively) compared to their common homozygotes.

Natriuretic peptide A (*NPPA*) rs5063 TT homozygotes were associated with having an edema recorded, although numbers affected were small (n=2/73, 2.74%, HR 4.40: 95%CI 1.09 to 17.76, *p*=0.04) compared to rs5063 common CC homozygotes with a rate of 0.84% (n=227/27,033).

*P3A4* rs2740574 CC homozygotes (n=4/26) had an increased risk for hospital diagnosed CKD compared to their common homozygotes groups (HR 1.14: 95% CI 1.01 to 1.29, *p*=0.04 and HR 3.01: 95%CI 1.13 to 8.02, *p*=0.03, respectively). *APCDD1* rs564991 CC homozygotes, *NOS1AP* rs10494366 heterozygotes (or GG+GT) and *NOS1AP* rs12143842 heterozygotes (or TT+TC) were less likely to have CKD compared to their common homozygotes.

## Discussion

Calcium Channel Blockers, especially dihydropyridines (dCCBs) such as amlodipine, are commonly prescribed to reduce raised blood pressures. Many pharmacogenetic variants have been reported as being of potential importance in dCCB responses based mainly on studies from laboratory studies, randomized trials or acute hospital settings, but data on clinical impact in routine care in the community have been limited. We estimated the association between 23 pharmacogenetic variants reported to affect dCCB response or adverse events in 32,360 patients prescribed dCCB using the UK Biobank-linked primary care data. Outcomes were assessed over a mean follow-up of over 10 years after first dCCB prescription. The most striking results were for the ryanodine receptor 3 (*RYR3)* rs877087, with T allele carriers having 13% increased risks of heart failure (p=0.02). In patients with a history of heart disease when first prescribed dCCB (N=2,296), *RYR3* homozygotes had 25% increased risk of heart disease diagnosis (MI, angina or heart failure) compared to CC variant (p*=*0.002). In addition, two genetic variants increased the likelihood of patients switching to an alternative antihypertensive medication (*NUMA1* and *CYP3A5*). The variant in *CYP3A5* also increased risk of CKD, and we hypothesise that it might be the reason for the switch in treatment, whilst variants in *ABCB1* and *APCDD1* increased risk of edema and CHD, respectively. Yet these variants were not associated with changes in medication. These adverse reactions are potentially preventable if patients were prescribed medications accounting for genotype. However, we found either little evidence of association with the studied adverse events for the majority of reported pharmacogenetic variants included, or a scattering of other associations, although some appeared genetically contradictory (with heterozygote and homozygote effects in opposite directions). Also, several variant associations had modest p-values that would be unlikely to survive reasonable multiple testing corrections.

*RYR3* mediates Ca2+ release from ryanodine-sensitive stores, triggering cardiac and skeletal muscles (35). A common variant in *RYR3* (rs877087) increased risk of HF in a study of 2,516 people randomized to amlodipine or to other antihypertensives (16). We support and extend this literature in a substantially larger sample using longitudinal analysis methods: we report increased HF risk in both TT homozygotes (n HFs=404 in 6,607 genotypes; HR 1.15) and CT heterozygotes (n HFs=943 in 15,377 genotypes; HR 1.12), compared to CC homozygotes (n=9,090). We used TWIST (36), a novel pharmacogenetic causal inference framework, to estimate the population average genetically modified treatment effect (GMTE) on HF if all *RYR3* T allele carriers could experience the same treatment effect as common CC homozygotes. We estimate their risk of HR would reduce by 9.2%. This corresponds to 170 avoidable HF diagnoses in the studied patients if they could be prescribed a hypertension medication not affected by *RYR3* rs877087. Furthermore, rs877087 has been associated with stroke in a genome-wide association study but the genotype effect in patients on treatment is unknown (25). Our findings suggest that rs877087 CT had an increased risk of hospital diagnosed stroke (n=203/15,381, HR=1.11, *p*=0.02) compared to common CC homozygotes, but we found no significant association when comparing TT to CC homozygotes.

Observational studies of drug effects often suffer from indication and other biases: as doctors aim to prescribe each medication based on the patients’ clinical state, statistically separating the effects of the medication from the effects of underlying disease is challenging, especially as data to correct potential confounders is seldom complete or entirely accurate. However, genotypes are inherited at conception and stay fixed, meaning that they pre-date receipt of the studied medications). Moreover, in the population under study, they do not directly influence treatment choice. Associations between genotypes and outcomes provide less confounded evidence than conventional observational associations, particularly because the participants and GPs were not told given genotype information by the UK Biobank study. Therefore, our finding that variants in two genes (*ABCB1* and *NPPA*) are associated with edema, and variants in two genes (*NUMA1* and *CYP3A5*) with switching treatment may result from dCCB pharmacokinetics or/and pharmacodynamics effects. A Korean study of 26 participants investigated the effect of *ABCB1* variants on pharmacokinetics of amlodipine (28) and found that rs2032582 CC carriers had decreased oral clearance of amlodipine, which might increase bioavailability (29) and eventually possible side effects. However, in our study the AA homozygote patients had increased risk of edema, but not the CC homozygotes.

*NUMA1* rs10898815 was previously identified in a GWAS of blood pressure (30) but as far we are aware there has been no reports on switching antihypertensive treatment. We found that AA homozygotes had increased likelihood of switching treatment, which was significant after multiple testing adjustment. CYP3A5, a cytochrome p450 enzyme, is a metaboliser of dCCB. *CYP3A5* *3 is the most common non-functional allele (rs776746-T) (with a prevalence of 6.6% in UK Biobank European cohort) which results in increased clearance of dCCB (31), resulting in less successful treatment. Our findings support this: TT (*CYP3A5*\*3) homozygotes had increased risk of CKD, for which high blood pressure is a risk factor, and were more likely to change treatments compared to common homozygotes (27 of 152 homozygotes switched vs. 3,420 of 28,184 homozygotes; HR 1.59, 95% CI 1.09 to 2.32, *p*=0.02).

In a pathway focused GWAS (32), genes in the *ADRA1* pathway ultimately affect intracellular calcium release (which dCCBs block) and blood pressure. The isoforms (i.e., a1A-adrenoceptor *(ADRA1A))* was associated with hypertension in patients. In a study in mice, it also mediated renal vasoconstriction in hypertension (33). Furthermore, *ADRA1A* affects renal functions via regulating Na+ reabsorption, renin secretion, renal blood flow and glomerular filtration rate, of which alterations cause kidney disease (33,34). Although we were not able to analyse GP recorded blood pressure measures robustly due to high rates of missing data, a SNP in *ADRA1A* (rs1048101) was associated with CKD. Patients homozygous for the common AA allele (30.4% of participants) had an increased risk of CKD, in contrast to GG homozygotes (20.3%) who had decreased risk. In TWIST analysis we estimated that 86 CKD diagnoses (7% of total) could be avoided if AA homozygotes could receive an alternative antihypertensive unaffected by the genotype.

In a previous GWAS the rs564991 C allele in *APCDD1* was associated with response to CCB (30). In our study we found that CC homozygotes had increased risk of MI/angina with a HR 1.12 (p=0.004); however, this was not significant after accounting for multiple testing.

Our analyses for edema were limited as only 1.02% of patients on dCCBs had GP recorded edema, substantially lower the reported prevalence of approximately 22% in the literature (7,8). This could be due to limitations in the data available; UK Biobank-linked primary care diagnoses include diagnostic codes only, with no free text. It has been discussed previously that estimates of the prevalence of edema depends on the study methods; in RCTs (randomised controlled trials) self-reported edema might be overestimated by the patients, or milder forms of edema might be reported compared to those that enter the GP record (8).

In routine clinical care patients’ blood pressure is regularly monitored after dCCB treatment initiation to determine whether the targets were achieved. However, we were unable to analyse this using UKB-linked primary care data due to the sparsity of blood pressure data available (only a small minority of patients had blood pressure in the record at the time of initiation, or within 2 months). This wide variation between patients in the time from prescription to next follow up meant we focussed instead on adverse drug reactions, not measured blood pressure.

The PharmGKB database (n=19 of the 23 studied variants were reported in PharmGKB) includes smaller and candidate studies, with only low or moderate levels of evidence for most of the relevant variants (as categorized by PharmGKB curators, often reflecting small sample sizes and lack of replication). Previously the most frequently studied genes were *CYP3A4* and *CYP3A5* with smaller sample sizes or non-European ancestry patients. The biggest study that we are aware of was a randomised control study 8,174 patients randomized to amlodipine (37). Additionally, a previous review about pharmacogenomics of hypertension medications reported 4 variants associated with dCCB response in a small Japanese sample (23). Of the 23 dCCB variants we found evidence for an effect on outcomes/adverse events for only 10 variants – even fewer after adjustment for multiple statistical testing - suggesting these few specific variants should be a priority for future study. The possible reasons for the lack of consistency include interethnic differences in studied populations, heterogeneity in exact phenotype studied, lack of adherence to medication, or variability in medication history between patients, but may also include publication biases, in which false positive statistical associations (type 1 errors) tend to be overrepresented, especially from small studies. Yet we here add substantially to the evidence base for these variants due in part to the large sample size studied, but also the strengths of analysing real-world primary care prescribing and the novel pharmacogenetic analysis approach triangulating evidence from multiple analysis methods (TWIST (36)). Using these data for pharmacogenetics analysis means we are able to look at more adverse reactions over longer periods and have therefore increased confidence for the relevance to routine clinical care of hypertension of variants where significant effects on outcomes are identified.

## Conclusions

Our analysis of longer-term prescribing in real-world primary care data support the hypothesis that use of genetic information in antihypertensive prescribing might optimize treatment selection for specific patients to maximize efficacy and reduce incidence of adverse events. The variants identified as associated with adverse clinical outcomes are good candidates for studies to test whether dCCB treatment outcomes can be improved with pharmacogenetic guided prescribing.

## Supporting information

dCCB Pharmacogenetics supplementary tables

## Data Availability

All data produced in the present work are contained in the manuscript

